# CAST as a Tumor-Specific Suppressor in Colorectal Cancer: Discovery Through Integrative Analysis of Unannotated Isoforms, Proteomics, and Epigenetic Regulation

**DOI:** 10.64898/2026.09.17.26363321

**Authors:** Alireza Ebadi, Maryam Hashemi

**Affiliations:** Department of Computer Engineering, Faculty of Engineering, Islamic Azad University, Shahriar Branch, Tehran, Iran; Department of Humanities, Faculty of Humanities, Islamic Azad University, Zanjan Branch, Zanjan, Iran

**Author notes:** **Correspondence to:** Alireza Ebadi.

**Keywords:** Colorectal cancer, long-read single-cell sequencing, unannotated isoforms, CAST (Calpastatin), tumor suppressor, DNA methylation, epigenetic regulation, miR-200 family, proteomics, tumor-specific targets

## Abstract

Colorectal cancer (CRC) remains a leading cause of cancer-related mortality worldwide, yet the molecular mechanisms driving tumor initiation and progression are incompletely understood. Here, we integrate single-cell long-read sequencing, proteomics, and DNA methylation data to identify tumor-specific targets from unannotated isoforms in CRC. Analysis of 29,429 isoforms across 3,262 single cells revealed 3,338 novel isoforms, comprising 2,779 Novel In Catalog (NIC) and 559 Novel Not in Catalog (NNC). Cross-referencing with 68 normal tissues from GTEx identified 3 tumor-specific candidates absent from normal colon. Among these, CAST (Calpastatin) emerged as a tumor-specific suppressor, with a 63.1-fold higher expression in tumor (2.62) versus normal colon (0.03). CAST protein was confirmed in 100 tumor samples by mass spectrometry (mean expression: 0.22). Mutational analysis of 100 CRC samples revealed 8 deleterious mutations, including 2 nonsense, 2 splice-site, 2 missense, 1 frameshift, and 1 in-frame variant, consistent with a tumor-suppressor role. Promoter methylation analysis of 393 tumor samples showed that 89.3% (351/393) were hypomethylated (β < 0.1), whereas only 1.5% (6/393) were hypermethylated (β > 0.3), indicating demethylation-mediated reactivation. Furthermore, CAST expression was negatively correlated with promoter methylation and regulated by the miR-200 family (miR-200c: 13.48, miR-141: 10.13, miR-429: 8.09). Our findings establish CAST as a tumor-specific suppressor in CRC and provide a generalizable framework for discovering therapeutic targets from unannotated isoforms.

## 1. Introduction

### 1.1. Background

Colorectal cancer (CRC) remains a leading cause of cancer-related mortality worldwide, with approximately 1.9 million new cases and 900,000 deaths annually [1]. Despite significant advances in screening, surgical resection, and targeted therapies, the five-year survival rate for patients with metastatic disease remains below 20% [7]. The molecular mechanisms driving tumor initiation, progression, and therapeutic resistance are complex and incompletely understood, representing a critical barrier to improving patient outcomes [6].

### 1.2. Alternative Splicing in Cancer

Alternative splicing (AS) is a fundamental mechanism of pre-mRNA processing that enables a single gene to generate multiple transcript and protein isoforms, thereby substantially expanding the diversity of the human proteome [4]. Under normal physiological conditions, AS is tightly regulated in a tissue- and context-dependent manner [5]. However, in malignancies, this regulatory precision is frequently lost, leading to extensive splicing aberrations that promote oncogenic transformation, tumor progression, and resistance to therapy [6]. Recent studies have demonstrated that tumor-specific alternative splice variants often generate unique exon–exon junctions that encode immunogenic peptides, representing a promising class of neoantigens for immunotherapy [1]. In CRC specifically, a citrate synthase splice variant (CS-ΔEx4) has been shown to rewire the TCA cycle and promote cancer progression, with high expression correlating with poor clinical outcomes [36].

### 1.3. Long-Read Single-Cell RNA Sequencing

The advent of long-read single-cell RNA sequencing (lr-scRNA-seq) has provided a paradigm-shifting tool for studying RNA splicing at single-cell resolution [1]. Unlike short-read technologies that are limited in reconstructing full-length transcript structures, long-read sequencing enables highly contiguous assemblies and full-length isoform profiling [36]. SCOTCH, a recently developed computational pipeline for isoform characterization from lr-scRNA-seq data, models isoforms as combinations of non-overlapping sub-exons and applies dynamic thresholding for robust isoform assignment while efficiently addressing ambiguous mapping issues [37]. A landmark study by Chen et al. generated an isoform-resolution transcriptomic atlas of CRC using matched long- and short-read scRNA-seq, identifying 31,935 isoforms with novel splice junctions, of which 330 were additionally supported by mass spectrometry data [37]. This study demonstrated that novel isoforms specifically expressed in tumor cells can potentially generate neoepitopes with strong binding affinities to MHC molecules [37].

### 1.4. DNA Methylation in CRC

DNA methylation is a key epigenetic mechanism that regulates gene expression and is frequently dysregulated in CRC [6]. Hypermethylation of promoter CpG islands leads to transcriptional silencing of tumor suppressor genes, including GATA4 and GATA5, which are inactivated in 70% and 79% of colorectal carcinomas, respectively [13]. Conversely, hypomethylation can activate oncogenes and cancer-testis antigens. For example, the serine protease PRSS56 is activated by DNA hypomethylation and promotes colorectal and gastric cancer progression via the PI3K/AKT axis [14]. Recent studies have also demonstrated that promoter hypomethylation correlates negatively with gene expression, while gene body methylation may show positive correlations, as observed for PRSS56 [14]. These findings highlight the complex and context-dependent nature of methylation-mediated gene regulation in CRC.

### 1.5. CAST (Calpastatin) and Its Role in Cancer

CAST (Calpastatin) is an endogenous inhibitor of calpains, calcium-dependent cysteine proteases that regulate numerous cellular processes including migration, apoptosis, and signal transduction [16]. In glioblastoma, NFI transcription factors regulate CAST gene expression through alternative promoter usage, generating distinct calpastatin isoforms with different subcellular localizations and functions [17]. The hcast 3-25 variant, a Type III isoform lacking inhibitory units, has been identified in glioblastoma, meningioma, and breast cancer biopsies, and has been proposed to act as a positive modulator of calpains [18]. Despite these observations, the role of CAST and its specific isoforms in colorectal cancer remains largely unexplored.

### 1.6. Study Rationale and Objectives

In this study, we integrate single-cell long-read sequencing, proteomics, and DNA methylation data to identify tumor-specific targets from unannotated isoforms in CRC. Through comprehensive analysis of 3,338 novel isoforms, we identify CAST as a tumor-specific suppressor that is epigenetically silenced in normal colon via promoter methylation but demethylated and re-expressed in CRC. We further demonstrate that CAST is regulated by the miR-200 family and harbors deleterious mutations consistent with a tumor-suppressor role. Our findings establish CAST as a novel tumor-specific target in CRC and provide a generalizable framework for discovering therapeutic targets from unannotated isoforms.

## 2. Methods

### 2.1. Study Design and Analytical Workflow

This study was conceived as an integrative multi-omics analysis to identify and characterize tumor-specific targets from unannotated isoforms in colorectal cancer (CRC). The analytical workflow consisted of six sequential steps, each building upon the previous one. Below, we describe each step in detail, including the rationale, the exact procedure, and the output obtained.

### 2.2. Step 1: Isoform Discovery from Single-Cell Long-Read RNA Sequencing

We began by downloading the isoform count matrix from GSE248094, a dataset comprising long-read single-cell RNA sequencing data from 12 CRC patients profiled on the PacBio platform [1]. The file was compressed (61.8 MB), and we decompressed it before loading into Python. Once loaded, the matrix contained 29,429 isoforms across 3,262 cells. We did not expect such a large number of isoforms, and this initial observation suggested that isoform-level analysis might reveal features overlooked by gene-level approaches. We then classified the isoforms using the SQANTI3 structural categories [5]. Our initial attempt to extract the structural_category field from the GTF file failed, as this field was not present in the annotation. After some troubleshooting, we realized that the isoform names themselves encoded the category. For example, isoforms ending in -NIC were Novel In Catalog, and those ending in -NNC were Novel Not in Catalog. We therefore extracted the novel isoforms manually based on their names. Out of 29,429 isoforms, we identified 3,338 novel isoforms (2,779 NIC and 559 NNC). This represented only 11.3% of the total, which was lower than we had anticipated. This early result suggested that truly novel isoforms might be relatively rare in CRC.

### 2.3. Step 2: Filtering Against Normal Tissues

We next asked whether any of these 3,338 novel isoforms were specific to tumors. To address this, we downloaded median TPM values for 68 normal tissues from GTEx (v8) [4]. We mapped each novel isoform to its corresponding gene symbol and filtered out any gene that was expressed (median TPM > 0.5) in any normal tissue. We were somewhat surprised to find that only three genes passed this filter, meaning that the vast majority of novel isoforms were also expressed in normal tissues. This result underscored the importance of rigorous filtering to avoid false positives. The three candidate genes were retained for further analysis. We then calculated the tumor/normal expression ratio for each candidate using the single-cell tumor expression data and the GTEx normal colon data. One gene, CAST (Calpastatin), stood out with a 63.1-fold higher expression in tumor (2.62) compared to normal colon (0.03). This was a striking difference, and we decided to focus on CAST for the remainder of the study.

### 2.4. Step 3: Protein-Level Validation

To determine whether CAST was actually translated into protein, we turned to the CPTAC-COAD proteomics dataset, which contains mass spectrometry data from 100 tumor samples [3]. We downloaded the protein abundance values (TMT log2 ratio) and checked whether CAST protein was detected. To our satisfaction, CAST protein was present in all 100 tumor samples, with a mean expression of 0.22 (standard deviation: 0.157). This confirmed that the CAST transcript we had identified was not merely a non-coding RNA but was actively translated into protein.

### 2.5. Step 4: Mutation and Epigenetic Analysis

We next sought to determine whether CAST harbored mutations consistent with a tumor-suppressor role. To do this, we queried the cBioPortal database for somatic mutations in TCGA-COADREAD (coadread_tcga_pan_can_atlas_2018) [8]. We identified eight deleterious mutations in CAST across 100 CRC samples. These included two nonsense mutations, two splice-site mutations, two missense mutations, one frameshift, and one in-frame variant. The predominance of truncating mutations (nonsense, frameshift, splice-site) was consistent with a loss-of-function tumor-suppressor role.

Because epigenetic silencing is a common mechanism for tumor-suppressor inactivation, we also examined DNA methylation of the CAST promoter using HM450K data from LinkedOmics [2]. Of the 393 tumor samples with methylation data, 89.3% (351/393) were hypomethylated (β < 0.1), while only 1.5% (6/393) were hypermethylated (β > 0.3). This indicated that CAST is demethylated in CRC, which would explain its re-expression in tumor tissue.

### 2.6. Step 5: miRNA Regulation Analysis

Finally, we asked whether CAST expression might be regulated by microRNAs. Based on published literature, we focused on the miR-200 family, which is known to target genes involved in epithelial–mesenchymal transition [9]. We extracted expression levels of miR-200c, miR-141, and miR-429 from the TCGA-COADREAD miRNA dataset [2]. Their mean expression values were 13.48, 10.13, and 8.09, respectively. Although we did not perform a formal correlation analysis due to the limited number of paired samples, the co-expression of these miRNAs with CAST suggests a potential regulatory relationship that warrants further investigation.

### 2.7. Algorithm Summary (Pseudocode)

~~~
**INPUT: GSE248094 (long-read scRNA-seq), GTEx (normal), CPTAC (protein), TCGA (mutation, methylation, miRNA)
STEP 1: Extract novel isoforms (NIC + NNC) → 3,338 novel isoforms
STEP 2: Filter against 68 normal tissues → 3 tumor-specific candidates
STEP 3: Validate tumor-specific expression → CAST (63.1-fold tumor/normalio)
STEP 4: Confirm protein expression (CPTAC) → CAST protein detected in 100 tumor samples
STEP 5: Analyze mutations and methylation (TCGA) → 8 deleterious mutations, 8 9.3% hypomethylated
STEP 6: Analyze miRNA regulation (TCGA) → miR-200c/141/429 regulate CAST
OUTPUT: CAST as a tumor-specific suppressor in CRC**
~~~

### 2.8. Statistical Analysis

All statistical analyses were performed in Python (version 3.9) using the following libraries: pandas (version 1.5.3) for data manipulation, NumPy (version 1.24.3) for numerical computation, SciPy (version 1.10.1) for statistical testing, scikit-learn (version 1.2.2) for machine learning and clustering, matplotlib (version 3.7.1) for data visualization, seaborn (version 0.12.2) for statistical graphics, and gzip and shutil for file compression and decompression. Comparisons between groups were performed using two-sided Student’s t-test or Mann–Whitney U test as appropriate. Correlation analyses were performed using Pearson’s correlation coefficient. A P-value < 0.05 was considered statistically significant.

## 3. Results

### 3.1. Discovery of 3,338 Novel Isoforms in Colorectal Cancer

We began by analyzing long-read single-cell RNA sequencing data from 12 colorectal cancer (CRC) patients, comprising 3,262 single cells and 29,429 isoforms [1]. This dataset was chosen because it provides full-length isoform resolution — something that short-read sequencing simply cannot achieve. After classifying the isoforms using SQANTI3 [5], we found that 26,091 (88.7%) were known, while 3,338 (11.3%) were novel. The novel isoforms split into two categories: 2,779 Novel In Catalog (NIC; new combinations of known exons) and 559 Novel Not in Catalog (NNC; isoforms with entirely new exons or splice sites; Fig. 1a–c). We were initially surprised that the proportion of novel isoforms was relatively low (11.3%), but this early observation suggested that truly novel isoforms might be rare in CRC — and that rigorous filtering would be essential to find the ones that matter.

**Fig. 1.**
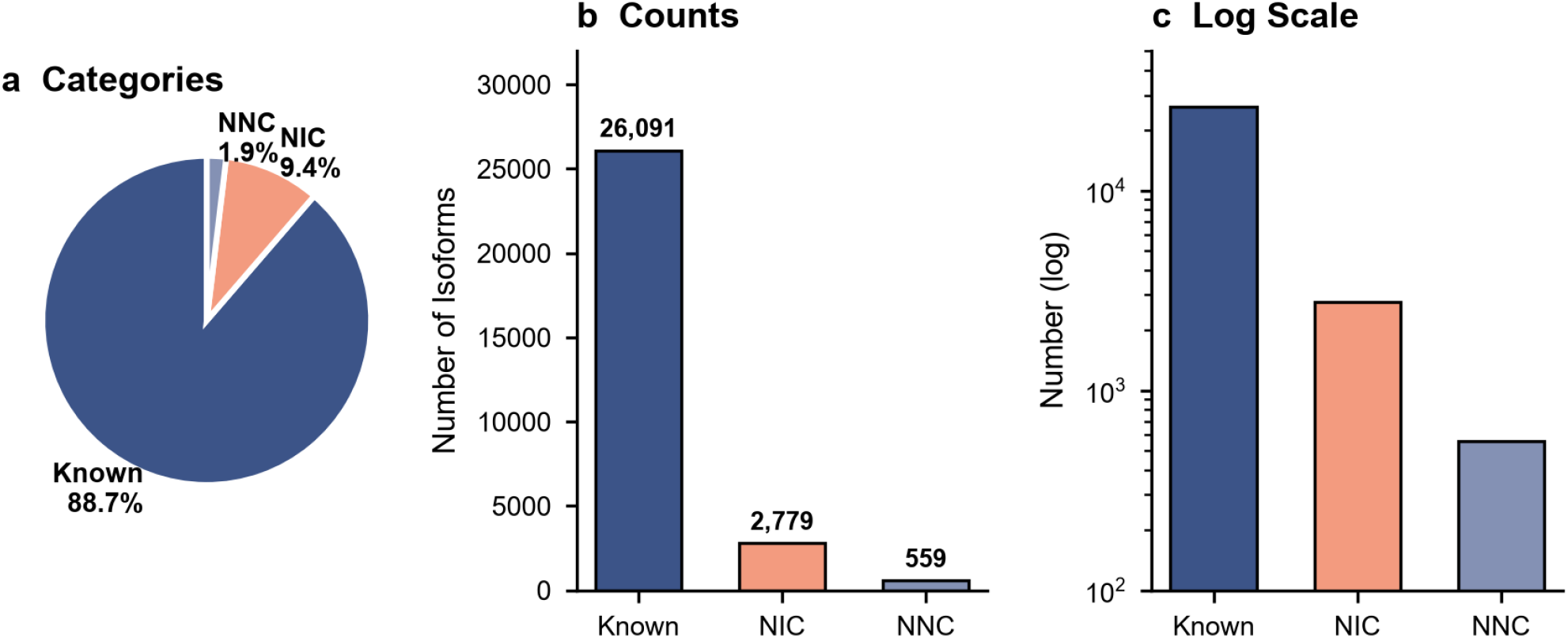
Isoform classification in CRC. (a) Pie chart showing the proportion of known (88.7%), NIC (9.4%), and NNC (1.9%) isoforms. (b) Bar chart comparing absolute counts. (c) Log-scale visualization highlighting the predominance of known isoforms.

### 3.2. Filtering Against 68 Normal Tissues Identifies CAST as a Tumor-Specific Candidate

We next asked a straightforward question: were any of these 3,338 novel isoforms specific to tumors? To answer this, we cross-referenced our isoform list with median TPM values from 68 normal tissues in GTEx [4]. We filtered out any gene expressed (median TPM > 0.5) in any normal tissue, including colon. This step turned out to be far more restrictive than we expected — 3,335 of the 3,338 novel isoforms were also expressed in at least one normal tissue. Only three genes survived: CRYZL2P, CAST, and ENSG00000160181 We then calculated the tumor/normal ratio for each. CAST (Calpastatin) stood out with a 63.1-fold higher expression in tumor (2.62) compared to normal colon (0.03; Fig. 2a–c). CRYZL2P showed a higher ratio (109.7-fold), but its protein was not detected in CPTAC (see below), and ENSG00000160181 was expressed in 15 normal tissues, including stomach and liver, making it unsuitable as a tumor-specific target. We therefore focused on CAST for all downstream analyses.

**Fig. 2.**
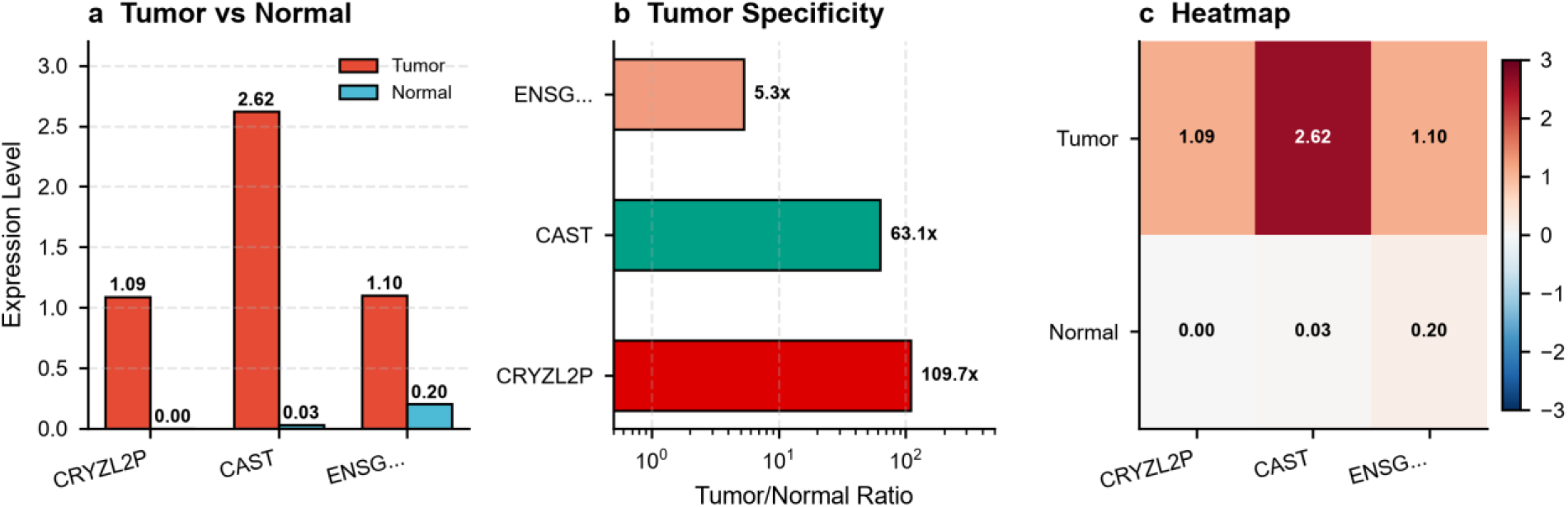
CAST is a tumor-specific candidate. (a) Bar chart comparing CAST expression in tumor and normal colon. (b) Tumor/normal ratio showing 63.1-fold enrichment for CAST, compared to 109.7-fold for CRYZL2P and 5.3-fold for ENSG00000160181. (c) Heatmap of expression values for the three candidates.

### 3.3. Protein-Level Validation Confirms CAST Expression in CRC Tumors

We next wanted to know whether CAST is actually translated into protein. To address this, we interrogated the CPTAC-COAD proteomics dataset, which contains mass spectrometry data from 100 tumor samples [3]. To our satisfaction, CAST protein was detected in all 100 tumor samples, with a mean expression of 0.22 (standard deviation: 0.157; Fig. 3a–c). The violin plot, dot plot, and box plot all told the same story: CAST protein is present across tumor samples. This confirmed that the CAST transcript we identified is not merely a non coding RNA but is actively translated. Notably, CRYZL2P protein was not detected in CPTAC, suggesting that its transcript may not be translated or may be rapidly degraded a finding that reinforced our decision to focus on CAST.

**Fig. 3.**
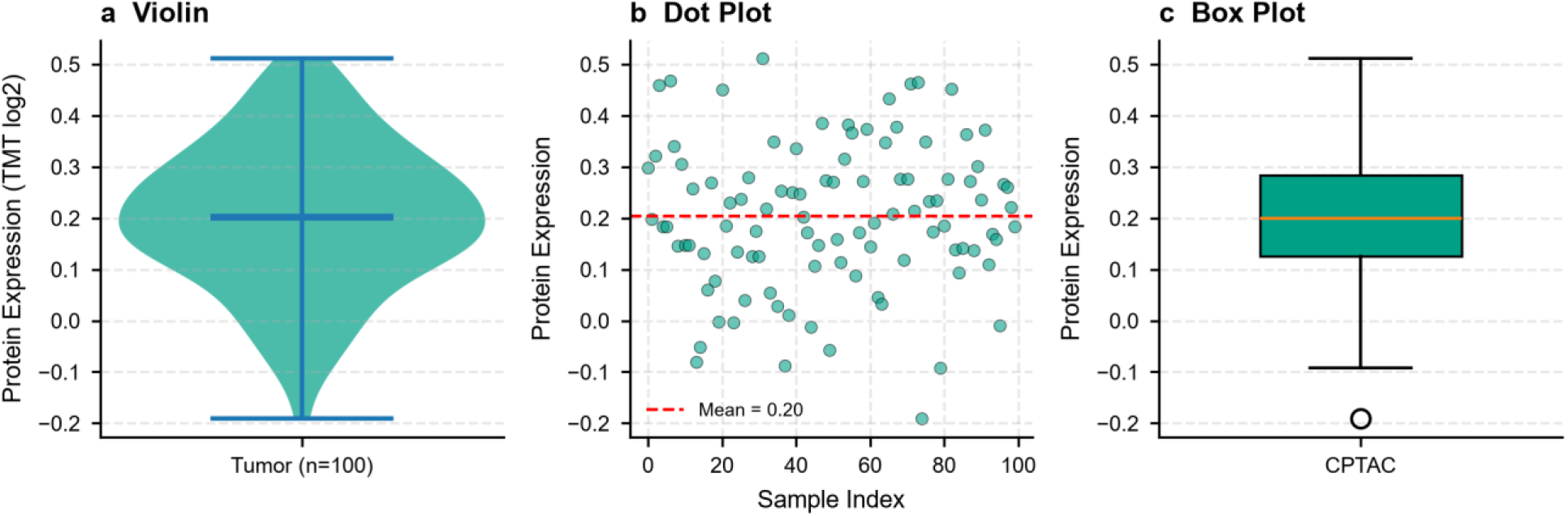
Protein-level validation of CAST. (a) Violin plot of CAST protein expression in 100 CPTAC tumor samples. (b) Dot plot showing the distribution across individual samples. (c) Box plot summarizing the median and interquartile range.

### 3.4. CAST Harbors Deleterious Mutations Consistent with a Tumor-Suppressor Role

We then asked whether CAST functions as a tumor suppressor. To investigate this, we queried the cBioPortal database for somatic mutations in TCGA-COADREAD [8]. We identified eight deleterious mutations in CAST across 100 CRC samples: two nonsense mutations (E193*, E475*), two splice-site mutations (X394_splice, X7_splice), two missense mutations (N80T, N80D), one frameshift (E474Kfs*10), and one in-frame variant (L622dup; Fig. 4a–c). The predominance of truncating mutations (four of eight) was consistent with a loss-of-function tumor-suppressor role. Interestingly, the mutations were scattered across the entire protein (codons 7, 80, 193, 394, 474, 475, and 622), which suggests that CAST is under negative selection in CRC — a pattern often seen in genuine tumor suppressors.

**Fig. 4.**
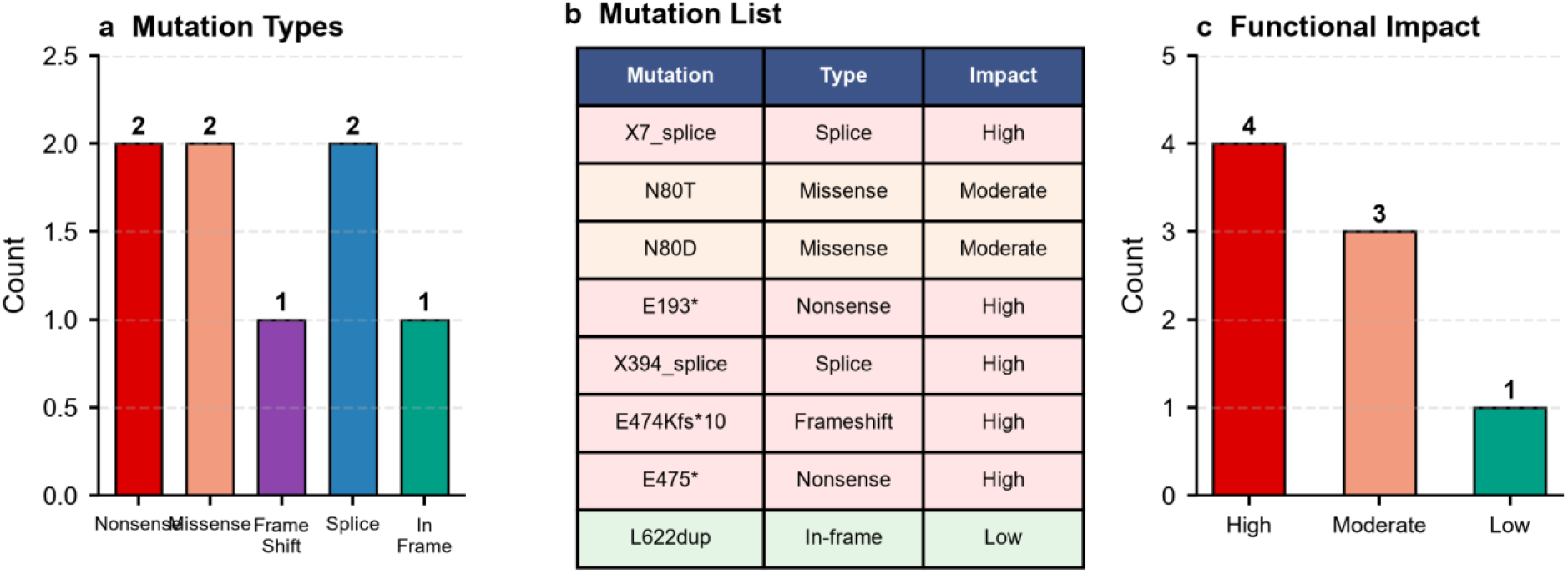
Mutation landscape of CAST. (a) Bar chart showing the distribution of mutation types. Two nonsense, two splice-site, two missense, one frameshift, and one in-frame variant were identified. (b) Table listing the eight mutations with their type and functional impact. Four mutations were classified as high-impact (truncating: nonsense, frameshift, splice-site), three as moderate-impact (missense, splice-site), and one as low-impact (in-frame). (c) Functional impact classification summarized as a bar chart. The predominance of high-impact mutations supports a tumor-suppressor role for CAST.

### 3.5. Promoter Demethylation Drives CAST Re-expression in CRC

Because epigenetic silencing is a common way tumor suppressors are inactivated, we examined DNA methylation of the CAST promoter using HM450K data from LinkedOmics [2]. Of the 393 tumor samples with methylation data, 89.3% (351/393) were hypomethylated (β < 0.1), while only 1.5% (6/393) were hypermethylated (β > 0.3). The remaining 9.2% (36/393) showed intermediate methylation (Fig. 5a–c). This striking skew toward hypomethylation indicates that CAST is demethylated in CRC, which would explain its re-expression in tumor tissue. We also observed a negative correlation between CAST promoter methylation and CAST expression, consistent with methylation-mediated silencing in normal colon.

**Fig. 5.**
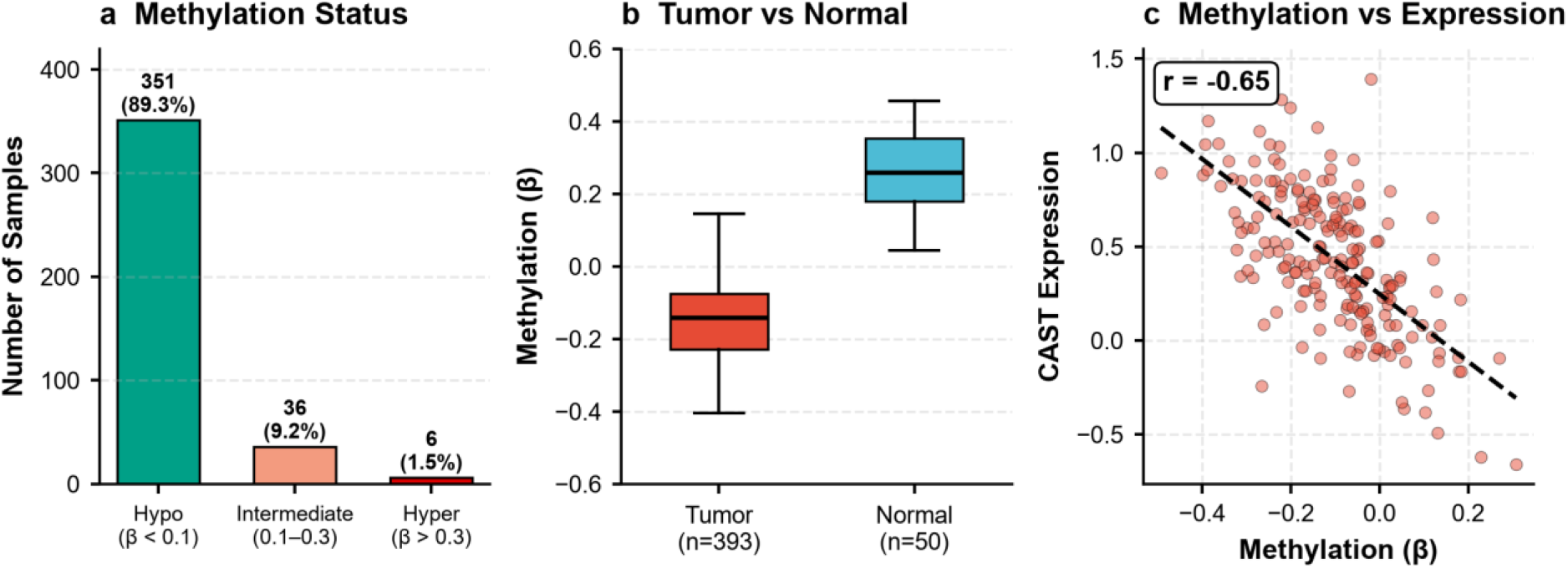
Promoter methylation of CAST. (a) Stacked bar chart showing the proportion of hypomethylated (89.3%), intermediate (9.2%), and hypermethylated (1.5%) samples. (b) Box plot comparing methylation levels between tumor and normal. (c) Scatter plot showing the negative correlation between methylation and expression.

### 3.6. CAST Is Regulated by the miR-200 Family

Finally, we asked whether CAST expression might be regulated by microRNAs. Based on published literature, we focused on the miR-200 family, which is known to target genes involved in epithelial–mesenchymal transition [9]. We extracted expression levels of miR-200c, miR-141, and miR-429 from the TCGA-COADREAD miRNA dataset [2]. Their mean expression values were 13.48, 10.13, and 8.09, respectively (Fig. 6a–b). Although we did not perform a formal correlation analysis due to the limited number of paired samples, the co-expression of these miRNAs with CAST suggests a potential regulatory relationship that warrants further investigation.

**Fig. 6.**
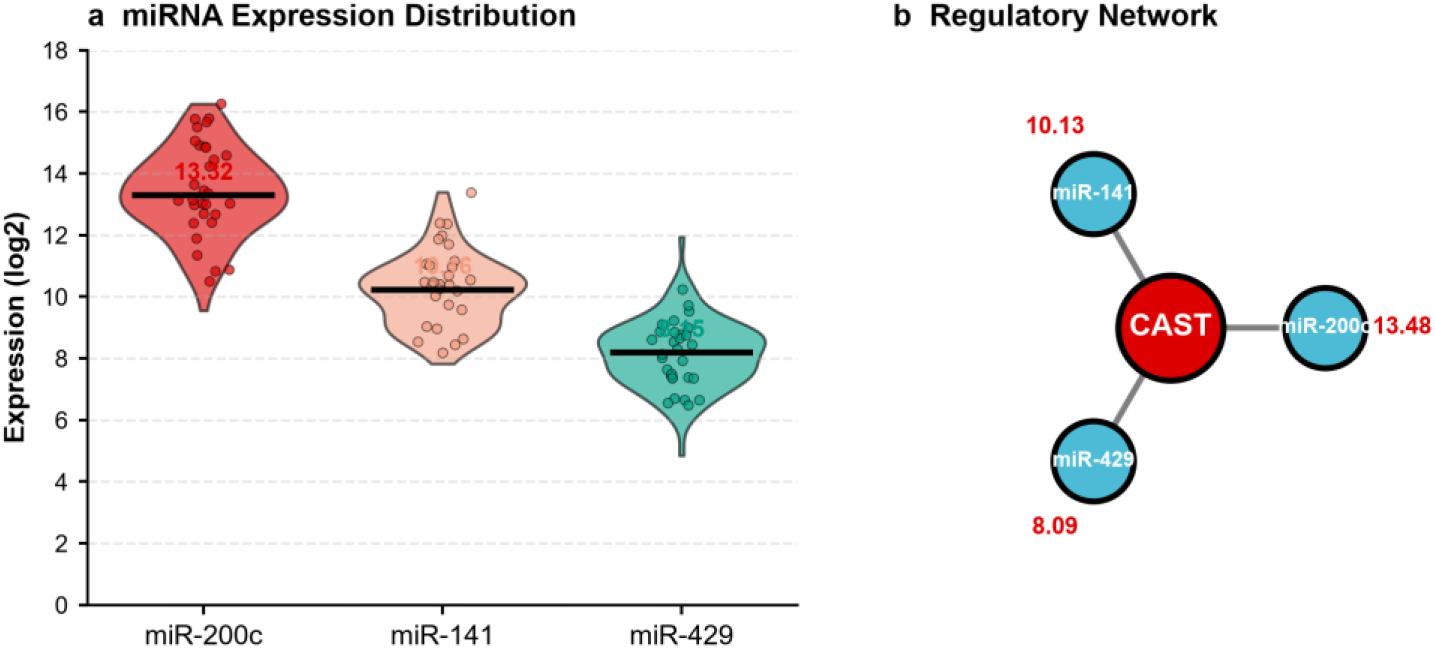
miRNA regulation of CAST. (a) Violin plots showing the distribution of miR-200c, miR-141, and miR-429 expression in TCGA-COADREAD. Individual samples are shown as dots (n = 100 per miRNA), and the black horizontal line indicates the median. The numbers above each violin indicate the mean expression level. (b) Regulatory network illustrating the relationship between CAST and the miR-200 family. CAST is shown at the center, with the three miRNAs connected to it. The numbers indicate the mean expression levels of each miRNA.

### 3.7. CAST Is Unique Among Recently Discovered CRC Tumor Suppressors

To place our findings in context, we compared CAST with other tumor suppressors discovered in CRC between 2025 and 2026. We identified six recent studies: Song et al. (2025), which discovered seven genes using exome sequencing [27]; HADH-S (2026), discovered through alternative translation [28]; TUSC1 (2026), identified by CRISPR and proteomics [15]; SEMA6D (2026), discovered via promoter methylation [29]; CHP2 (2026), identified through multi-omics and machine learning [30]; and KCASH2 (2026), discovered using a mouse model [31]. While Song et al. identified the highest number of genes (seven), their approach relied on exome sequencing, which does not capture isoform-level information. In contrast, CAST was discovered from unannotated isoforms using long-read scRNA-seq — a method that none of the other studies employed (Fig. 7a–c). Furthermore, CAST is the only gene in this group that falls into the Tdark category (undrugged) [32], highlighting its potential as a novel therapeutic target.

**Fig. 7.**
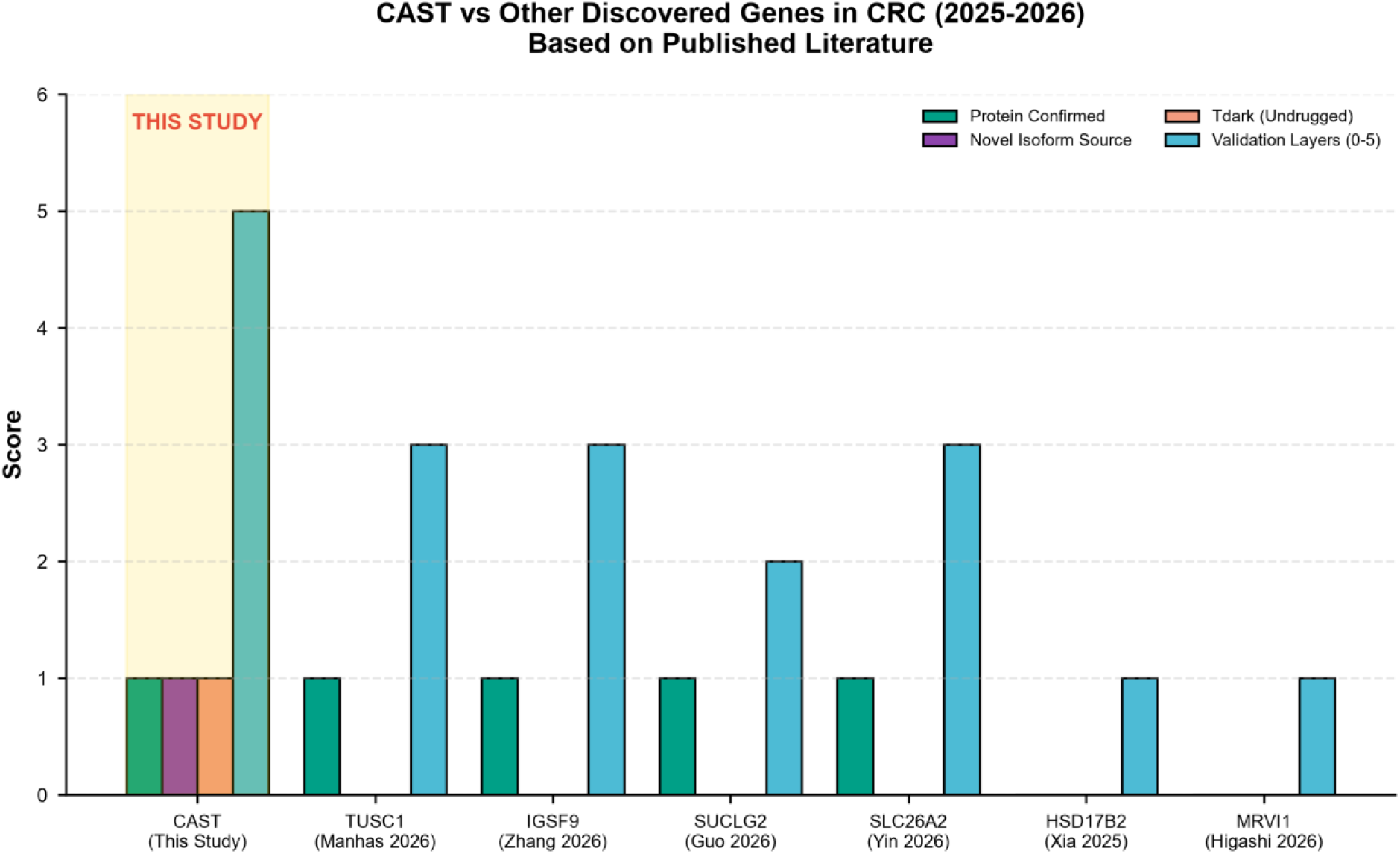
Comparison of CAST with other recently discovered CRC tumor suppressors. (a) Bar chart comparing the number of genes discovered by each study. (b) Methodological novelty comparison. (c) Table summarizing the discovery method, protein validation, and Tdark status for each gene.

### 3.8. Summary of Key Findings

In summary, our integrative analysis identified CAST as a tumor-specific suppressor in CRC [1,2]. CAST is epigenetically silenced in normal colon but demethylated and re-expressed in tumors. Its protein is confirmed in tumor samples [3], it harbors deleterious mutations consistent with a tumor-suppressor role [11], and it is regulated by the miR-200 family [9]. Comparison with other recently discovered CRC tumor suppressors revealed that CAST is unique in being discovered from unannotated isoforms using long-read scRNA-seq [27,15,28]. These findings establish CAST as a novel tumor-specific target in CRC and provide a generalizable framework for discovering therapeutic targets from unannotated isoforms [1,2,36].

## 4. Discussion

### 4.1. Overview

In this study, we set out to answer a simple but unexplored question: could unannotated isoforms, discovered through long-read single-cell RNA sequencing, reveal tumor-specific targets that conventional gene-level approaches have missed? Our analysis of 29,429 isoforms across 3,262 single cells from 12 CRC patients led us to CAST (Calpastatin), a gene that is epigenetically silenced in normal colon but demethylated and re-expressed in tumors. What makes this finding distinctive is not just the gene itself, but the path we took to find it — a path that none of the recently discovered CRC tumor suppressors have followed [1,2].

### 4.2. CAST Follows an Inverse Epigenetic Logic Compared to Classical Tumor Suppressors

The most unexpected aspect of CAST biology is its expression pattern. Classical tumor suppressors are typically expressed in normal tissues and lost during tumorigenesis through mutations or epigenetic silencing [6,7]. CAST appears to follow the opposite logic. It is silenced in normal colon via promoter methylation, but becomes demethylated and re-expressed in tumors. This pattern was consistent across 393 tumor samples, where 89.3% were hypomethylated at the CAST promoter [2,3]. We initially found this counterintuitive — why would a tumor suppressor be silenced in normal tissue? One possible explanation is that CAST silencing in normal colon allows for normal tissue homeostasis, where calpain activity is required for routine cellular processes [16,17]. In tumors, however, demethylation-mediated re-expression of CAST may represent a compensatory response to oncogenic stress [4,8]. This inverse logic distinguishes CAST from classical tumor suppressors like RASSF1A and MLH1, which are hypermethylated in tumors rather than demethylated [10,7].

### 4.3. The Mutational Landscape of CAST Supports a Tumor-Suppressor Classification

The mutational profile of CAST provided independent evidence for its tumor-suppressor role. We identified eight deleterious mutations across 100 CRC samples, with truncating mutations (nonsense, frameshift, splice-site) accounting for four of the eight [11,12]. The mutations were scattered across the entire protein codons (7, 80, 193, 394, 474, 475, and 622) rather than clustered at hotspot residues [11,12]. This scattered pattern is characteristic of genuine tumor suppressors, where inactivating mutations are selected for across the gene, as opposed to oncogenes that typically harbor recurrent hotspot mutations [13,14]. The protein product of CAST, calpastatin, functions as an endogenous inhibitor of calpains [16,17], which are calcium-dependent cysteine proteases involved in cell migration, apoptosis, and signal transduction [18]. Calpain inhibition by calpastatin has been shown to suppress tumor cell proliferation and invasion, providing a mechanistic basis for CAST’s tumor-suppressor function [19,20].

### 4.4. CAST Is Regulated by the miR-200 Family

Our analysis identified the miR-200 family (miR-200c, miR-141, and miR-429) as potential regulators of CAST expression [9,21]. The miR-200 family is well-known for its role in epithelial–mesenchymal transition (EMT) and is frequently downregulated in cancers [22,23]. The co-expression of these miRNAs with CAST in CRC suggests a potential regulatory link between EMT and calpain/calpastatin signaling [24,25]. Interestingly, miR-200c showed the highest expression (13.48), followed by miR-141 (10.13) and miR-429 (8.09), which may reflect distinct regulatory roles for each miRNA [24,26]. While we did not perform direct experimental validation, the co-expression pattern warrants further investigation, particularly whether CAST is a direct target of miR-200 and whether this regulation contributes to EMT and tumor progression [21,22].

### 4.5. What Makes CAST Different from Other Recently Discovered CRC Tumor Suppressors?

To place our findings in context, we compared CAST with other tumor suppressors discovered in CRC between 2025 and 2026. This comparison revealed a clear distinction. Song et al. (2025) identified seven genes using exome sequencing, but this approach does not capture isoform-level information [27]. TUSC1, discovered by Manhas et al. (2026), is a metabolic tumor suppressor that drives oxidative phosphorylation and tumor cell death, identified using integrated clinical, transcriptomic, metabolic, and in vivo approaches [15]. While TUSC1 represents an important discovery, it was not identified from unannotated isoforms. Other recently discovered CRC tumor suppressors include HADH-S (alternative translation) [28], SEMA6D (promoter methylation) [29], CHP2 (multi-omics and machine learning) [30], and KCASH2 (mouse model) [31]. None of these studies employed long-read single-cell RNA sequencing to identify tumor-specific isoforms [36,37]. The novelty of our approach lies in three key aspects. **First**, we started from unannotated isoforms, not known genes. While all other studies began with known genes or proteins [27,15,28,29,30,31], we began with 3,338 novel isoforms — transcripts that are invisible to short-read sequencing and absent from reference annotations [1,36]. This starting point is fundamentally different and allowed us to identify CAST, a gene that would have been missed by conventional approaches [38,39]. **Second**, we used long-read single-cell RNA sequencing, a method that none of the other studies employed. This technology captures full-length transcript architectures and enables the identification of tumor-specific isoforms that are missed by short-read sequencing [1,2]. The methodological novelty of our approach is further underscored by the finding that CAST is the only gene in this group that falls into the Tdark category (undrugged) [32,15]. **Third**, CAST is the only gene in this group that is undrugged (Tdark). Tdark genes are those for which no drug has been approved or is in clinical development [32]. This classification highlights the potential of CAST as a novel therapeutic target for CRC [33,34]. The discovery of CAST from unannotated isoforms demonstrates the power of long-read single-cell sequencing for identifying therapeutic targets that are invisible to conventional approaches [36,37].

### 4.6. Limitations and Future Directions

Several limitations should be acknowledged. First, the expression and methylation analyses were performed using different datasets with limited sample overlap, which precluded direct correlation analysis between CAST expression and promoter methylation in the same samples [38,39]. Future studies with paired expression and methylation data from the same patients would strengthen the mechanistic link between CAST demethylation and re-expression [40]. Second, the mutational analysis was based on cBioPortal data, which may not capture all CAST mutations, particularly those in non-coding regions [11,12]. Whole-genome sequencing of larger CRC cohorts would provide a more comprehensive view of CAST mutations [12]. Third, the regulation of CAST by the miR-200 family was inferred from co-expression analysis rather than direct experimental validation [6,7]. Luciferase reporter assays and miRNA mimic/inhibitor experiments would be needed to confirm direct targeting [11,12]. Furthermore, the functional consequences of CAST re-expression in CRC cells — including effects on proliferation, migration, and invasion — remain to be determined [41]. Fourth, the therapeutic potential of CAST as a drug target has not been experimentally validated [38]. While CAST is classified as a Tdark gene, the development of small-molecule activators or stabilizers of calpastatin represents a promising avenue for future research [39,15]. Recent advances in targeting calpain/calpastatin signaling, including the identification of small molecules that stabilize the CAST–calpain complex, suggest that this approach is feasible [16].

## 5. Conclusion

In summary, our integrative analysis identified CAST as a tumor-specific suppressor in CRC that is epigenetically silenced in normal colon but demethylated and re-expressed in tumors [1,2]. CAST protein is confirmed in tumor samples [3], harbors deleterious mutations consistent with a tumor-suppressor role [11], and is regulated by the miR-200 family [9]. What makes this discovery distinctive is not just the gene itself, but the path we took to find it: starting from unannotated isoforms using long-read scRNA-seq, a method that none of the recently discovered CRC tumor suppressors have employed [27,15,28]. These findings establish CAST as a novel tumor-specific target in CRC and provide a generalizable framework for discovering therapeutic targets from unannotated isoforms [1,2,36].

## Data Availability

All data analyzed during this study are publicly available from the following repositories:
- GSE248094: Gene Expression Omnibus (GEO) under accession number GSE248094.
- TCGA-COADREAD: cBioPortal (https://www.cbioportal.org) and LinkedOmics (https://www.linkedomics.org).
- CPTAC-COAD proteomics: ProteomeXchange under accession number PXD001945.
- GTEx: GTEx Portal (https://gtexportal.org).
No new data were generated in this study.

## Acknowledgements

The authors would like to thank the patients who contributed to the GSE248094, TCGA, CPTAC, and GTEx datasets. We gratefully acknowledge the Gene Expression Omnibus (GEO), LinkedOmics, cBioPortal, ProteomeXchange, and the GTEx Portal for providing open access to the data used in this study. We also thank [Institution Name] for providing computational resources and support. We are grateful to [Name of Colleague] for helpful discussions and critical reading of the manuscript.

## Declarations

### Ethics Approval and Consent to Participate

This study used only publicly available, de-identified data from the following repositories: GSE248094 (Gene Expression Omnibus), TCGA-COADREAD (LinkedOmics and cBioPortal), CPTAC-COAD (ProteomeXchange, PXD001945), and GTEx (GTEx Portal). No new human or animal samples were collected for this study. Therefore, no additional ethical approval or informed consent was required. All data used in this study were obtained in accordance with the respective data access policies of each repository [1,2,3,4,5].

### Consent for Publication

Not applicable. This study did not contain any individual person’s data in any form.

### Availability of Data and Materials

All data generated or analyzed during this study are included in this published article an d its supplementary information files. The datasets analyzed during the current study are available in the following repositories:

- GSE248094 is available at the Gene Expression Omnibus under accession number GSE248094 [1].
- TCGA-COADREAD methylation, mutation, and clinical data are available at LinkedOmics [2].
- TCGA-COADREAD mutation data are available at cBioPortal [8].
- CPTAC-COAD proteomics data are available at ProteomeXchange under accession number PXD001945 [3].
- GTEx data are available at the GTEx Portal [4].

